# COVID-19 self-testing in Nigeria: Stakeholders’ opinions and perspective on its value for case detection

**DOI:** 10.1101/2022.01.28.22269743

**Authors:** Veronica A. Undelikwo, Sonjelle Shilton, Morenike Oluwatoyin Folayan, Oluwatoyin Alaba, Elena Ivanova Reipold, Guillermo Z. Martínez-Pérez

**Affiliations:** Department of Sociology, University of Calabar, Calabar, Cross River State, Nigeria; FIND, the global alliance for diagnostics, Geneva, Switzerland; Department of Child Dental Health, Obafemi Awolowo University, Ile-Ife, Osun State, Nigeria; Institute of Public Health, Obafemi Awolowo University, Ile-Ife, Nigeria

**Keywords:** Nigeria, COVID-19, Community representatives, Self-testing, Diagnostics, Qualitative research

## Abstract

**Background:** COVID-19 testing coverage is limited in Nigeria. Access to SARS-CoV-2 self-testing kits may help improve the detection of asymptomatic and mildly symptomatic cases and increase the currently low rate of COVID-19 testing in the country. Before implementing SARS-CoV-2 self-testing in Nigeria, it is imperative to assess the populations’ perceptions regarding this innovation. We therefore conducted a qualitative study to investigate people’s values and preferences for SARS-CoV-2 self-testing in Nigeria.

**Methods:** We used semi-structured interviews and focus group discussions among healthcare workers, community representatives, and public health implementors to explore values and perceptions around various aspects of COVID-19 testing, including conventional COVID-19 testing, SARS-CoV-2 self-testing, the safe and effective use of SARS-CoV-2 self-testing, actions upon receiving a positive SARS-CoV-2 self-test result, and future prospects for SARS-CoV-2 self-testing.

**Results:** Respondents reported that there is limited availability of conventional SARS-CoV-2 testing in Nigeria. While just a few respondents were familiar with SARS-CoV-2 testing, respondents generally supported the use of SARS-CoV-2 self-testing as they felt it could assist with early case detection and improve access to testing. Concerns relating to the use of SARS-CoV-2 self-testing were majorly about the ability among low literacy populations to use and interpret the test, the affordability of tests, equity of access, and the availability of healthcare system support for those who test positive.

**Conclusion:** Though the public perceive multiple benefits associated with access to SARS-CoV-2 self-testing, the efficiency of the national health service delivery system may limit access of the users of the kits to psychosocial and clinical support. In Nigeria, where COVID-19 vaccine coverage is low and the risk of further waves of COVID-19 is high, self-testing may assist in the prompt detection of cases and contribute to halting the spread of the virus.

## 1 Introduction

Coronavirus disease 2019 (COVID-19) is a novel disease that has caused a global pandemic, resulting in more than 332 million infections and 5.5 million deaths in January 2022 (1). It is an airborne, respiratory infection that is easily transmissible between individuals. Although vaccines against COVID-19 can reduce the severity of infection, they do not eliminate the risk of infection or transmission of the infection (2). There is a need for sustainable COVID-19 containment strategies to halt its transmission, especially in low- and middle-income countries (LMICs), where COVID-19 vaccine coverage remains low and the risk of multiple further waves of the pandemic is high (3).

One effective strategy to help contain COVID-19 is community-wide testing to enable prompt detection of cases. The most accurate technology for the detection of SARS-CoV-2, real-time reverse transcription polymerase chain reaction (RT-PCR), can determine whether a person is currently infected with SARS-CoV-2 (4). However, LMICs have few RT-PCR-equipped laboratories, and limited resources to provide essential reagents (5). To facilitate community-level case identification, rapid antigen tests, in the form of lateral flow assays, represent a low-cost, portable, and easy-to-perform solution for LMICs, although they are less sensitive than RT-PCR. Multiple asymptomatic cases of COVID-19 may go undetected (6). To reduce this risk, self-tests for serial or frequent home-use enable people to test self-collected specimens and detect SARS-CoV-2 infection without the direct assistance of healthcare professionals (7, 8). While not yet widely introduced in most LMICs, the commercialization and distribution of SARS-CoV-2 self-tests has already been approved across Canada (8), the United States (9), and India (10).

In some LMICs, self-administered rapid HIV, malaria, and syphilis tests are already widely used (11-15). The World Health Organization (WHO) has recently released recommendations for hepatitis C self-testing (16). The acceptability of self-testing among the general population is usually high, as these approaches can help to ensure higher levels of confidentiality, they are usually more affordable and accessible, and they guarantee freedom of choice of testing location (13, 17). As with other self-testing devices, self-testing for SARS-CoV-2 may be a feasible solution to resource-constrained governments’ lack of capacity to carry out mass screening for COVID-19, provided there are clear pathways to ensure self-testing users can access treatment and can isolate when needed.

In Nigeria, the country in West Africa worst affected by the COVID-19 pandemic (18), the concept of individuals having access to technologies for self-testing for infectious diseases is not new. The acceptability of HIV self-testing is high (19-21), although its use is not yet widespread, and so is the acceptability of malaria self-testing by the general public (22) and healthcare workers (23). Access to SARS-CoV-2 self-testing kits may help increase the prompt detection of infection in asymptomatic and mildly symptomatic cases and improve the currently low rate of COVID-19 testing in the country (24). It could also reduce the resistance to seeking care that results from the stigma associated with COVID-19 infection (25).

To develop and issue recommendations for regulatory and public health practice around SARS-CoV-2 self-testing in Nigeria, it is imperative to conduct a thorough assessment of the population’s perceptions regarding the innovation, as this can provide insights into socio-culturally acceptable strategies for implementing self-testing, helping to address any barriers and accelerating its widespread use. We therefore conducted a qualitative research study to investigate the values and preferences of the general population around SARS-CoV-2 self-testing in Nigeria.

## 2 Methods

### 1.1 Study Design and Site

For this qualitative inquiry we used semi-structured interviews (SSIs) and focus group discussions (FGDs). The study was conducted in Nigeria by the Institute of Public Health, Obafemi Awolowo University, Ile-Ife with the support of FIND. This was an ancillary study to a larger, population-based survey conducted in Nigeria between July and September 2021, which assessed the general public’s values and acceptance around SARS-CoV-2 self-testing (hereafter referred to as “self-testing”) (26).

### 2.1 Population and Sampling

The study population comprised three groups of stakeholders who hold decision-making capacities for the future usage of self-testing. Healthcare workers (HCWs) were targeted because of their capacity to recommend (or not recommend) the use of self-testing to their patients. Representatives, or spokespersons, of various civil society communities (RCSs) were targeted because of their capacity to influence community decision-making on the utility of self-testing and guide people on what to do following a reactive self-test result. Potential COVID-19 self-testing implementers (PIs) were targeted because of their capacity to decide to pool financial and human resources to procure and distribute self-testing at scale, for example in the workplaces they managed or in the geographies where they had jurisdiction to regulate or operate. Common inclusion criteria for all populations were: aged 18 years or more, willing to provide informed consent, and fluent in English or could communicate in broken English.

Efforts were made to ensure maximum variation in sampling in terms of gender, urban and rural workplaces, and professional and institutional profiling. To ensure a diversity of voices was represented in the sample, a purposive sampling approach was used. Sex-disaggregated lists of at least 50 profiles per study population were produced. To avoid sampling by convenience, these lists were randomly rearranged by FIND staff using the RANDOM.Org^®^ randomizer. The interviewers contacted potential informants by phone, starting with the first name on each list. Potential informants were provided with information about the study’s aim and procedures, and those who expressed an interest in participating were asked to partake in either an SSI or an FGD.

### 2.2 Data Collection and Processing

All informants gave their informed consent. Depending on the informants’ expressed preferences, data collection was conducted either using Zoom^®^ teleconferencing software or in-person at a designated place convenient for the informant and the interviewer. Each informant chose the language in which the interview was to be conducted.

The data collection was led by a team of research assistants with qualitative research experience. The same 45-item structured guide was used for SSIs and FGDs. The guide included questions around six main topics: knowledge of conventional COVID-19 testing; values around self-testing; the public’s preferences for the delivery of self-testing; safe and effective use of self-testing; actions taken upon receiving a reactive self-test result; and future prospects for the distribution of self-tests (26). The interviewers posed the 45 questions to the informants in the same order and probed them further depending on the nature of their responses.

All encounters were audio-recorded. Zoom^®^ encounters were not video-recorded. The recordings were transcribed verbatim into MS-Word^®^ files. Responses not in English were translated into English. All transcripts and translated sections within the transcripts were cross-checked by the analysts (VAU, OA, MOF) against the recordings, for accuracy and completeness.

### 2.3 Data Analysis

Transcripts were uploaded into Quirkos^®^ software, and a thematic comparative analysis was applied. First, all transcripts were deductively coded using a pre-defined coding scheme (26). Whenever an emerging theme was identified, new codes were inductively created. In parallel with the coding, the analysts prepared reflexive memos to control for the risk of informant bias.

Iteratively with the coding, the dataset was analyzed using a four-stage approach: Transcript by transcript at first; followed by a theme-by-theme, sex-sensitive comparison of coded narratives across all transcripts and then by a theme-by-theme rural versus urban-sensitive comparison of coded narratives across all transcripts; and finishing with a trans-study population comparison of key findings.

The reports were prepared taking into consideration general insights as well as insights from isolated or deviant cases. The informants’ own words were used to prepare the report. Attention was paid to the memos to ensure that no analysts’ informant biases were being introduced. The COREQ guidelines were considered.

### 2.4 Ethics Approval

This study received ethics approval from the Health Research Ethics Committee of the Obafemi Awolowo University in Ile-Ife (Ref. IPH/OAU/12/1730). All informants signed an informed consent form and received a copy. Prior to any data collection, the informed consent forms were shared by email with the respective informants to give them more time to make an informed decision about their participation. Participants who attended the in-person FGDs were compensated for their transportation costs. As per criteria set during the informed consent process, the transcripts of participants’ encounters with the interviewers were not shared with any person outside of the research team.

## 2 Results

### 2.5 Participants’ Characteristics

Two FGDs and ten SSIs were conducted with each of the three study populations. On average, the FGDs and SSIs lasted for 55 minutes and 122 minutes, respectively. A total of 58 informants (29 female) participated (Supplementary Material 1). Half of the informants were either living and/or working in rural Osun State. The mean age of informants was 45 years. Most participants (55) had completed tertiary education (diploma, bachelors, or masters). Among the 19 HCWs, 5 were nurses. There was diversity in terms of the institutional representation of PIs and RCSs. To protect their anonymity, demographic information highlighted in Supplementary Material 1 only indicates their socio-professional sector of influence.

The findings are presented as per the four core themes included in the analysis process, namely: uptake of conventional COVID-19 testing; values around SARS-CoV-2 self-testing; safe and effective use of self-testing; and future prospects for the delivery of self-testing. Unless otherwise specified, the voices reported below were common across the three study populations.

### 2.6 Uptake of Current COVID-19 Testing Modalities

Access to conventional facility-based testing was described as being of most interest to travelers and symptomatic patients. Testing was not considered to be in great demand for case detection among mildly symptomatic people. Several deterrents to testing for the general public were identified, including the high cost, frequent delays to receive test results, a generalized perception that COVID-19 was low-risk, and a fear of isolation and being “stigmatized”. Walk-in visits to health facilities to demand testing by members of the community was described as limited because communities were perceived as either “poorly educated” about COVID-19 symptoms, lacking in “motivation” to request tests, or were unable to afford them.

> *It has to do with the early stigmatization. Once someone is tested positive to COVID-19, the society and even the immediate family discriminates against him, and this has been a contributory factor for discouraging people to go and test*. (SSI 26, rural male PI)

Among other reasons given for the low demand for testing was the suggestion that “disbelief” about COVID-19 was commonplace and that there were misconceptions about COVID-19 being synonymous with malaria:

> *Many people still believe that COVID does not exist, that is just like malaria and that they don’t have to go for testing because if they are being diagnosed of COVID: that maybe is a death sentence, that they have to isolate them. As you know, isolation is like you are taking them away from their family, from their home*. (SSI 19, urban female HCW)

HCWs noted that the detection of early infections is difficult as most people present to health facilities at an advanced stage of the disease. Additionally, all study groups perceived there to be a dearth of clinic-or laboratory-based testing sites. Most HCWs expressed that they were not involved with COVID-19 testing and emphasized that the scarcity of diagnostic centers, together with facility staff being too busy caring for patients, limit the healthcare system’s capacity for community-based case detection. The shortage of trained professionals to conduct COVID-19 testing, poor availability of COVID-19 diagnostics, and limited access to personal protective equipment were other limiting factors for the routine testing of symptomatic patients and their contacts. The RCSs also mentioned language barriers, lack of privacy, poor safety, and low wages as barriers for healthcare workers to conduct community-based testing.

All informants who had direct (e.g., collection of nasal or blood samples) or indirect (e.g., being a member of the State’s COVID-19 committee) experience of testing resided or worked in an urban area. None of the informants from rural areas reported any type of experience with COVID-19 testing and hence, as many of them reflected, lacked factual knowledge in relation to testing sites, techniques, and operators. While the HCWs were aware that COVID-19 could be diagnosed using rapid antigen testing, many RCSs and PIs could not explain in any detail what diagnostic technologies for COVID-19 were available in their contexts.

### 2.7 Value of SARS-CoV-2 Self-testing

Of all the informants, just three HCWs were aware of self-testing. They had learned about it through the social media, CNN and Al Jazeera, and an international journal. Despite the general lack of knowledge around self-testing, most informants had an opinion on its potential advantages. Self-testing was perceived to be an innovation that would help end-users reduce costs, time, and other resources necessary to access COVID-19 diagnostic centers. It was defined as a potentially private, convenient, and easy way to obtain a prompt diagnosis of SARS-CoV-2 infection, to facilitate access to early treatment and, as a consequence, to reduce COVID-19-attributable mortality. It was also noted, especially by the HCWs, that the workload and “stress” among health facility personnel would be reduced. Special emphasis was placed on the assertion that the confidentiality of self-testing results would help some end-users overcome their fear of stigma:

> *One advantage is it will make the detection of the disease very easy, because it could actually serve as a facilitator because people will prefer to do the test themselves in the comfort of their homes instead of going out to health facility and then everybody starts looking at them and thinking that “does this person have COVID-19 or not?”* (SSI 5, rural female RCS)

Some potential disadvantages, more related to the idea of self-testing among specific end-users than to the technology itself, were also identified. As per the informants’ narratives, some end-users, especially populations with low levels of literacy, may be less able to correctly interpret the results. The possibility of obtaining invalid results due to poor compliance with the test’s instructions was also frequently mentioned. It was suggested that some end-users may self-medicate or may deny a positive result and thus refuse to seek medical treatment.

Although there was consensus that the availability of self-testing in Nigeria may improve public interest in COVID-19 testing, it was also suggested that it would be mainly travelers who may prefer to use self-testing to avoid the “stress of doing a PCR”, that the “elites” would be among the first to use them as they have more information and resources to obtain them, and that urban dwellers will show more interest in self-testing than their rural counterparts. The HCWs further perceived that various cadres of healthcare professionals may themselves benefit from the regular use of self-testing if they are exposed to COVID-19 in the workplace.

The majority of informants stated they would recommend self-testing as they considered it could lead to early commencement of treatment for those who might need it, and of measures to avoid further transmission of the virus. Some HCWs also expressed that they would be keen to recommend self-testing to their communities, as this could help alleviate their daily workload in healthcare facilities.

The informants’ likelihood of recommending self-testing, however, might also be influenced by factors such as price, ease of use, availability, and accuracy. To ensure ease of use, step-by-step instructions for the use of self-test kits should be provided in English, Igbo, Hausa, and Yoruba. Some RCSs and PIs noted that user instructions should also be provided in Braille. Information suggested for inclusion in the instructions included: how to unpack, use, and dispose of the kit; how to read and interpret the result; what the time interval before a repeat test should be; and what to do if the result is positive. As one PI elaborated, if the kits are designed with full consideration of the country’s low-literacy levels, most self-test end-users will be able to perform the test, in the same way that diabetes patients with low literacy levels are able to use their glucose monitoring devices:

> *Glucostix is there and it is graded in different color codes. That is the sort of thing to be done, so that even an illiterate, someone who is not educated, know that the moment you see red, it means danger. So, you don’t need to put figures there. You can use color codes*. (SSI 22, urban female PI)

To tackle the likely barrier of unaffordability for a large proportion of the Nigerian populace, a few PIs and all RCSs opined that self-testing kits should be delivered free-of-charge. Conversely, some HCWs, PIs, and RCSs opposed the free distribution of kits on the premise that the public “do not value what is free”. If the devices had to have a market price, the preferred maximum cost expressed by RCSs and PIs was Naira (N) 250 and N500, respectively (N100 is approximately US$0.25). HCWs held the most varied views, with some suggesting pricing ranging between N100 and N500 and others suggesting pricing ranging between N1000 and N2500.

Regarding availability, it was suggested that a range of stakeholders from the public (e.g., healthcare workers), private not-for-profit (e.g., non-governmental organizations, NGOs; civil society organizations, CSOs), and private for-profit (e.g., pharmacies, patent medicine vendors) sectors should be engaged with the distribution of the kits. As per the informants’ suggestions, kits could be made available in hospitals, churches, mosques, football fields, cinemas, barbing salons, or through NGO/CSO community and house-to-house outreach programs.

With regard to accuracy, and as claimed by some of the HCWs partaking in the FGDs, their likelihood to recommend self-testing would be conditional on the kits clearly indicating that they had been approved by the National Agency for Food and Drug Administration and Control (NAFDAC, see: https://www.nafdac.gov.ng/). The public’s preferred test specimens would be sputum, urine, and saliva; blood collection was considered to be too invasive, as it would require a professional to perform it and was thus the least preferred specimen.

> *People are beginning to clamor for non-invasive procedures. I would love a situation whereby the use of saliva can be explored. Everybody spits all over the place, so we shouldn’t*… *Now what we are doing is a throat swab and everything, but if you have done that testing… you would know that “oh my God!” Especially the nasal one, it’s painful*. (SSI 22, female urban PI)

### 2.8 Safe and Effective Use of COVID-19 Self-Testing

While some RCSs and HCWs opined that there were no circumstances under which access to self-testing should be restricted, others challenged this perspective. Some PIs were of the opinion that minors should have limited access to self-testing. An urban, female PI thought that access to self-testing should be limited when there is “no longer an upsurge in infection rates” and the perception of risk associated with COVID-19 is low. Some RCSs expressed that self-testing should be restricted if the distributors start “hoarding the kit among themselves” or if there is any security risk such as “kidnapping (of people distributing the kits)”. Some HCWs added that, to avoid misinterpretation of results or use of expired kits, elderly persons living alone and individuals of any age with limited literacy should have limited access to self-testing.

Although all informant groups were clear that an indicator of success of self-testing could be end-users’ communication of their results to health authorities, all groups insisted that a fear of death, isolation, and stigma were reasons for potential under-reporting. The HCWs also noted that people’s concerns about health facility-induced stress, resulting from being passed through multiple departments to receive COVID-19 care, could also be a driver of under-reporting.

It was noted that isolation for those who receive a positive self-test result might be feasible for the “elites”. Isolation was perceived to be dreaded by most Nigerians and especially by those of “low socio-economic status who live in crowded spaces”. For many families it is simply impossible to isolate for 10 to 14 days unless they receive support from an NGO/CSO or, as some PIs emphasized, direct financial support. Despite isolation being a measure recommended by health authorities, the HCWs expressed empathy and understanding of the reasons why members of the public might not comply with this.

> *They may wish to isolate but circumstances may not allow them. Like, if they are sharing rooms with members of their family, if they are not living in personal environment, they may not be able to isolate. So, the only thing they can do is for them to just protect themselves, or use their face mask, and they should ensure the people around them use their face mask*. (FGD 4 with urban HCWs)

Individuals’ non-compliance with isolation following a positive COVID-19 result was not the biggest concern for many of the informants. Some HCWs thought that although some people may use self-testing and refuse to disclose a positive result, there was still a likelihood that they would take all possible precautions not to infect others. Other RCSs and PIs opined that some end-users would not report a positive result to a healthcare facility because they would want to manage the disease themselves. One RCS noted that he would simply communicate at work that he was “ill” and would also warn and protect his family members, but he would not report his COVID-19 status.

> *Some [people] working with private organization don’t want to take permission to be off work because stigmatization is there too. Once they hear that you are positive for whatever, they will ask you to “Just stay at home and don’t even bother to come back again”*.(FGD 4 with urban HCWs)

Individuals who perceive that a positive result “means death” may be at risk of psychosocial ill health, while “resilient” individuals may be more likely to react in way that protects others. The impact of a positive result on an individual will depend on their “personality”, level of education, and location of residence. The breadwinners in a household and people with co-morbidities might be particularly concerned about the impact receiving a positive test result might have. To some informants, the “common man” does not perceive COVID-19 to be “fatalistic”, and it is mostly the “elites” that are more afraid. Women were thought to be able to react more positively to a positive result than men and young people who, as per some informants’ opinions, have generally poorer health-seeking behaviors than women.

Irrespective of personal attributes, most informants believed that many individuals might be “psychologically disturbed” after receiving a positive result. The impact might manifest in the form of avoiding people, not going out, becoming “depressed”, suffering from insomnia, losing the ability to concentrate, or feeling “lonely” and “afraid of the unknown”.

> *The person is going to test himself or herself, and then of course [is going to] know the result alone, which gives some confidentiality. However, the disadvantage is that it can lead to some mental issues, like depression and possibly suicidal tendency if not properly managed*. (SSI 23 with urban male PI)

There was consensus that a supportive environment may mitigate this impact. If end-users received pre- and post-test counseling, they would be “psychologically prepared” for a positive result. A few HCWs suggested that end-users be counseled on the use of a self-test before receiving it. All groups stressed the need for sustained public education and sensitization through outreach activities and seminars carried out through churches, mosques, social media, and television and radio broadcasts. NGOs/CSOs could play a key role in the dissemination of information at a community-level. A key action to mitigate the risk of psychosocial harm would be to make clear to end-users, in the kits’ written instructions, that effective linkage to COVID-19 care will occur should they receive a positive self-test result.

### 2.9 Future Prospects for SARS-CoV-2 Self-Testing

All groups expressed the opinion that treatment provision and contact-tracing following an end-user self-reporting in a clinic might be difficult due to a lack of adequate human and logistical resources. To prevent the public becoming disappointed with self-testing, the health sector must be strengthened by increasing the number of staff in healthcare facilities to cater for the volume of clients who might attend for the management of a COVID-19 infection following self-testing. Other steps should include improving health facilities’ existing staff capacity to manage cases effectively, irrespective of their severity; providing personal protective equipment to all staff tasked with direct management of cases; and increasing the number of facilities closer to the community, where end-users could both report a positive result and receive clinical care. PIs and RCSs identified the need for closer collaboration between healthcare workers and the community, including community development workers, to ensure that users of self-tests receive an appropriate response.

Other barriers to be addressed prior to the distribution of self-tests included the possible inability of end-users to afford the kits; anticipated poor distribution and unequal accessibility to the kits throughout the country; poor awareness about the availability of kits; and the likelihood of “hoarding” or stock-outs of self-test kits. To address these barriers, it was proposed that the kits should be rendered affordable through government subsidy, accessible from medical supply outlets in all communities, and introduced to the population following provision of adequate public education.

To promote community uptake of self-testing, community mobilization could be sustained using both printed and web-based social media. The “fear of death” should not be used in promotional messages. Rather, public messaging should emphasize “responsibility to care”. Some RCSs suggested that acceptability among the public may improve if, during public education efforts around self-testing, the government does not promote any “insinuation” that efforts promoting COVID-19 self-testing are for “ulterior motives” (i.e., in reference to possible suspicions that government officials may be profiting from the introduction of self-testing in their communities). The PIs noted that uptake could be promoted if opportunities for vaccination and treatment were concurrently provided at self-testing distribution points, with simultaneous national policies mandating regular self-testing in work environments.

## 3 Discussion

This study harnessed the opinions of critical stakeholders who would be involved in the rollout of SARS-CoV-2 self-testing in Nigeria. These stakeholders included representatives of communities who might become the potential end-users of self-testing; healthcare workers who might advise community members on self-testing access and usage; and implementers from the private and public sectors who have access to resources, make decisions in relation to the rollout of self-testing, and are in a position to support the country’s continued access to self-testing kits and post-testing care. There was consensus across all three groups that self-testing would be of considerable value in helping to overcome some of the current individual-, health system-, and community-level barriers to ensure access to and benefit from conventional healthcare facility-based COVID-19 testing. Nevertheless, the uptake and use of self-testing was not perceived to be free of challenges. To overcome any potential risks associated with the misinterpretation of results, misuse of kits, or under-reporting of reactive results, the informants also proposed strategies to promote the uptake of self-testing in a viable way and to guarantee counseling and healthcare provision to those whose self-test result is positive for SARS-CoV-2.

One of the values of self-testing identified by our informants was that it may reduce the burden on overstretched healthcare facilities. Self-testing offers opportunities for asymptomatic individuals or those with a mild case of infection to either rule out the possibility of having a SARS-CoV-2 infection or to seek care only in the event of a reactive result. In any case, plans must be instituted to accommodate a likely increase in the number of self-testers that may visit their nearest clinic requesting confirmatory testing and specialist care. The risk of further burdening the healthcare system can be reduced if plans are made to scale-up facilities’ capacity to respond to any increase in self-test-diagnosed cases prior to any rollout of self-testing.

Some of the structural barriers identified for facility-based COVID-19 testing, such as the cost of healthcare, unavailability of diagnostics and therapies, and rejection of the “diseased” by certain sections of the public, might affect the uptake of self-testing if left unaddressed. The cost of healthcare already hampers the uptake of and adherence to HIV services (27) and preventive care (28), and it is a critical consideration for provision of laboratory services (29). The informants suggested that self-testing devices should be subsidized, although concern was expressed by some that the cost of isolation might be a greater worry than the cost of self-testing.

Concerns about “hoarding” and stock-outs were expressed in our study. For future implementation of self-testing, it will be important to identify which distribution and accountability models will be the most cost-effective in making self-testing available (and affordable) in areas where the communities have concerns regarding the governance of health product supplies. As hinted by some informants, NAFDAC could make a key contribution, by passing stringent regulations on self-test distribution and quality assurance, to mitigate the risk of unavailability of quality self-test kits throughout Nigeria.

In determining the most cost-effective models for the distribution of self-tests, other emotion-related factors interact with cost and the regulatory framework. The psycho-emotional burden of receiving a positive self-test result must also be considered. As with HIV infection, COVID-19 infection is associated with stigma (30), which implies that for distribution models to be cost-effective they must include provisions to mitigate the fear of being stigmatized for having COVID-19 and, as a consequence, incurring social and economic loss or deprivation. In the absence of provision of psychosocial support and clear pathways for linkage to post-self-test care, even the best distribution models may fail. Our study emphasizes the need for pre- and post-self-test counseling provision, as well as for the engagement of various stakeholders from the public and private not-for-profit healthcare provision sectors, to support provision outside of the regular healthcare system.

The stress associated with the possibility of isolation must be acknowledged and addressed as one of the most impactful impediments to testing as a whole, professional use or self-test. This is a reasonable concern for many men who are burdened with the need to provide care for their family as the sole bread winner in many households, as well as for many women who work in the informal economy and rely on their daily wages to provide for their children (31). In a country with sections of the population severely affected by high rates of malnutrition and extreme poverty, a debate is urgently needed on which measures would be the most effective, and acceptable to both society and health authorities, to ensure that people who are infected who cannot isolate will not transmit SARS-CoV-2 to others.

Isolation seemed to be a greater concern than fear of morbidity. This study did not provide an understanding on why there might be a low perception of risk (i.e., individuals’ judgments about and evaluations of hazards to which they may be exposed) for COVID-19 disease among some Nigerians though a prior study has made some suggestions. This low perception of risk is also a barrier to the use of self-testing. Nevertheless, what this study has identified is the need to tailor appropriate risk communication and education to enable individuals to understand their risks (32), when resorting to malaria treatment in the absence of either a malaria or a COVID-19 test, or self-managing a COVID-19 infection without having at least warned their relatives and other close contacts.

Gender norms are another structural factor that cannot be transformed in the short-term and that may affect self-testing usage. Self-testing distribution models must include targeted strategies to encourage the uptake of self-testing by men and adolescents who, as per our respondents’ voices, are perceived to exhibit limited use of health services or to be persons with worse healthcare behaviors than women. Lessons on entry strategies for self-testing may, therefore, be learned from the introduction of HIV self-testing that specifically targeted men (19).

Our study has some strengths and limitations that should be considered. The informants were recruited from both urban and rural areas of Nigeria, and diversity regarding gender identities, location of work, and socio-professional profiles was ensured. However, this was a qualitative study, and the informants’ insights may not be representative of all possible opinions in the country. Our findings offered themes and insights that might be characteristic of the specific groups represented in our sample. Additionally, some data collection encounters were carried out via Zoom^®^. The content of interviews conducted online and in-person was similar; however, the interviewers felt that it was easier to build rapport with the interviewees when partaking in face-to-face encounters. The possibility that informants interviewed via Zoom^®^ changed their narratives due to privacy or confidentiality concerns cannot be disregarded.

In conclusion, facilitating the adoption and use of self-testing in Nigeria will require multiple layers of planning, ranging from the active engagement of policymakers to develop regulations and strategies for the rollout of a national self-testing program, to capacity-building of health institutions to manage the increased demand that may result from the rollout, and to the active engagement of communities and community decision-making platforms to allay fears and to support and promote the effective use of self-testing. While the public may perceive that access to SARS-CoV-2 self-testing will be beneficial in the long-term, the structures and systems in health care institutions must be prepared to provide appropriate psychosocial and clinical support to self-testers. For a populous country like Nigeria, where COVID-19 vaccine coverage remains low and the risk of further epidemic waves of COVID-19 is looming, self-testing holds promise for allowing communities themselves to promptly detect cases and contribute to halting the spread of the virus in the region.

## Data Availability

Due to the nature of this research, participants of this study did not agree for their data to be shared publicly, so supporting data is not available.

## 5 Author Contributions

GZMP, SS, and EIR developed the initial research project. MOF adapted the research protocol and led the implementation of the study in Nigeria. VAU, OA, and MOF performed the data processing and analyses. VAU, GZMP, SS, and MOF wrote the manuscript. All authors have reviewed the final version of the manuscript.

## 7 Acknowledgments

The authors are greatly indebted to all informants who agreed to participate in this study and to share their insights with us. This article has been submitted as a pre-print on medRxiv.

## Supplementary Material

**Table 1:**
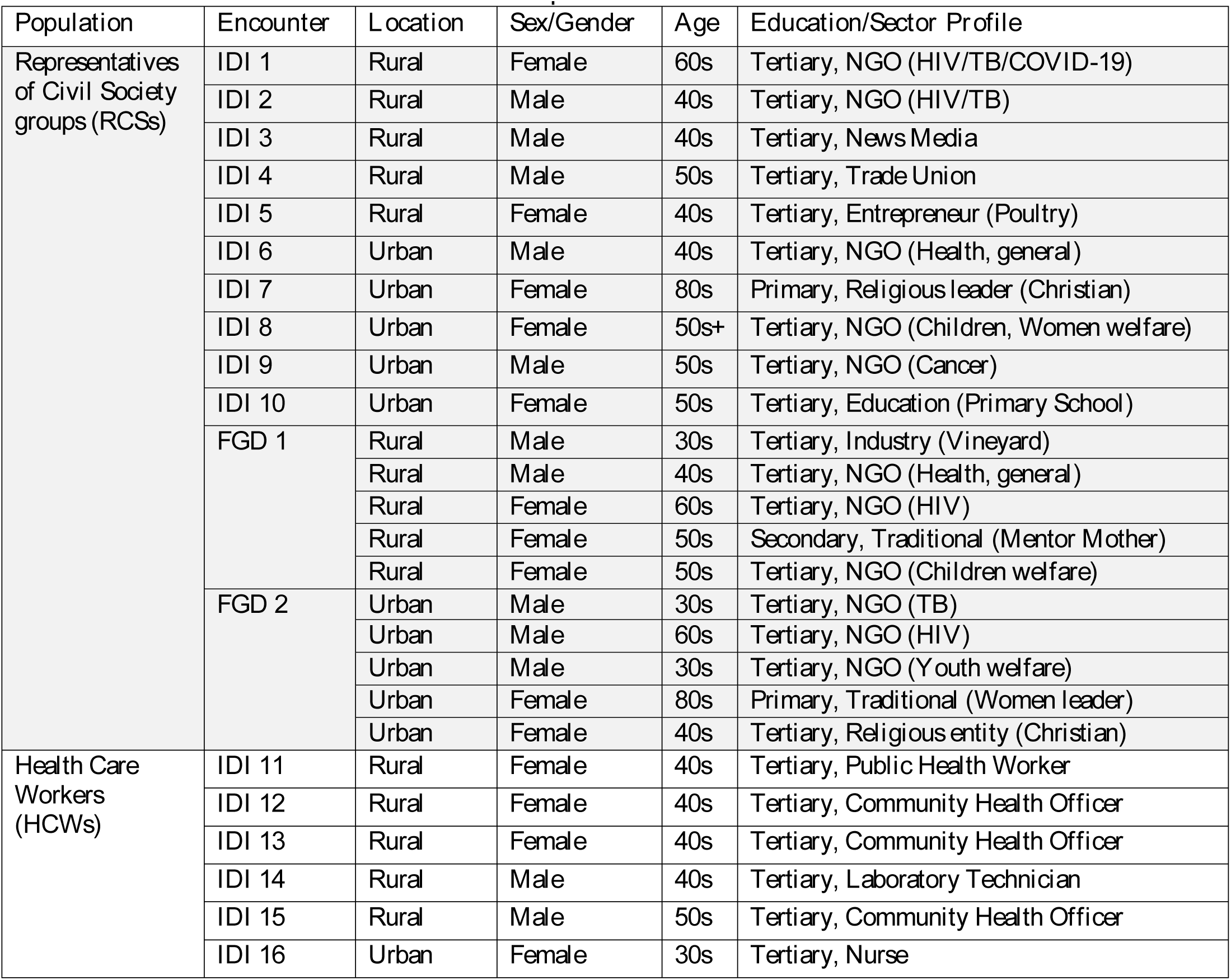
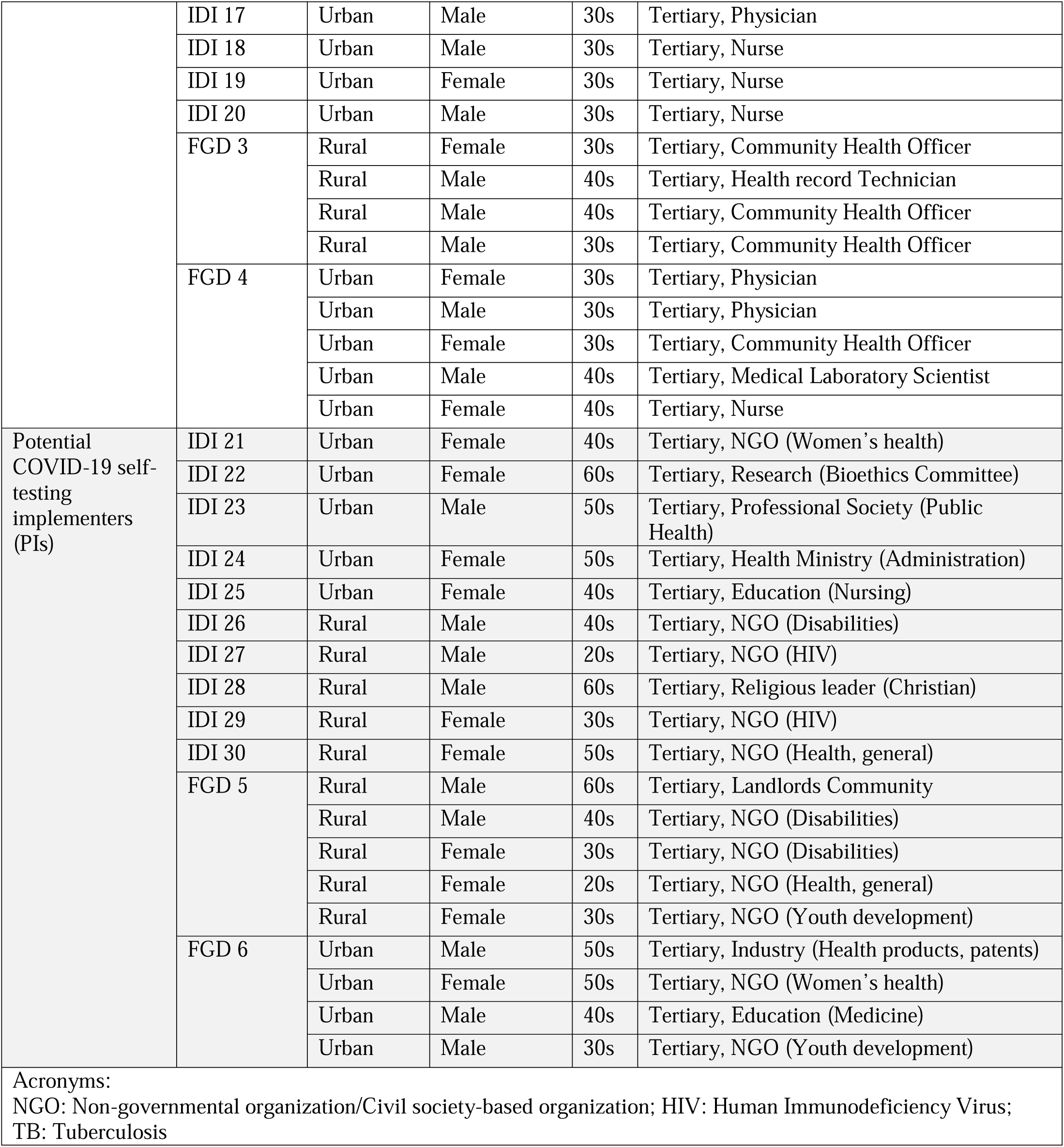
Participants’ characteristics.

## References

1. World Health Organization. Coronavirus disease (COVID-19) pandemic Geneva: World Health Organization; 2021 [Available from: https://www.who.int/emergencies/diseases/novel-coronavirus-2019?adgroupsurvey={adgroupsurvey}&gclid=Cj0KCQiA-eeMBhCpARIsAAZfxZAJ4dAylz4XOhRsUIEvbd7w347fha9E4SKI8GW3Ix8jZArMQOCkhwgaApItEALw_wcBItEALw_wcB.

2. CDC. Science Brief: COVID-19 Vaccines and Vaccination 2021 [Available from: https://www.cdc.gov/coronavirus/2019-ncov/science/science-briefs/fully-vaccinated-people.html.

3. World Health Organization. WHO Director-General’s opening remarks at the media briefing on COVID-19 – 11 March 2020 Geneva: World Health Organization; 2020 [Available from: https://www.who.int/director-general/speeches/detail/who-director-general-s-opening-remarks-at-the-media-briefing-on-covid-1911-march-2020.

4. Yüce M, Filiztekin E, Özkaya KG. COVID-19 diagnosis -A review of current methods. Biosens Bioelectron. 2021;172:112752.

5. Giri AK, Rana DR. Charting the challenges behind the testing of COVID-19 in developing countries: Nepal as a case study. Biosafety and Health. 2020;2(2):53–6.

6. Oran DP, Topol EJ. Prevalence of Asymptomatic SARS-CoV-2 Infection : A Narrative Review. Ann Intern Med. 2020;173(5):362–7.

7. Hengel B, Causer L, Matthews S, Smith K, Andrewartha K, Badman S, et al. A decentralised point-of-care testing model to address inequities in the COVID-19 response. Lancet Infect Dis. 2021;21(7):e183–e90.

8. Self-testing.gov.gr. Οδηγίες προς τους Πολίτες 2021 [Available from: https://self-testing.gov.gr/.

9. U.S. Food and Drug Administration. Coronavirus (COVID-19) Update: FDA Authorizes Antigen Test as First Over-the-Counter Fully At-Home Diagnostic Test for COVID-19 2020 [Available from: https://www.fda.gov/news-events/press-announcements/coronavirus-covid-19-update-fda-authorizes-antigen-test-first-over-counter-fully-home-diagnostic.

10. Indian Council of Medical Research. COVID-19 Home Testing using Rapid Antigen Tests (RATs) Indian Council of Medical Research; 2021 [Available from: https://www.icmr.gov.in/pdf/covid/kits/COVID_Home_Test_Kit_06012022.pdf.

11. Ibitoye M, Frasca T, Giguere R, Carballo-Diéguez A. Home testing past, present and future: lessons learned and implications for HIV home tests. AIDS Behav. 2014;18(5):933–49.

12. Kabaghe AN, Visser BJ, Spijker R, Phiri KS, Grobusch MP, van Vugt M. Health workers’ compliance to rapid diagnostic tests (RDTs) to guide malaria treatment: a systematic review and meta-analysis. Malar J. 2016;15:163.

13. Sy TRL, Padmawati RS, Baja ES, Ahmad RA. Acceptability and feasibility of delegating HIV counseling and testing for TB patients to community health workers in the Philippines: a mixed methods study. BMC Public Health. 2019;19(1):185.

14. Kpokiri EE, Marley G, Tang W, Fongwen N, Wu D, Berendes S, et al. Diagnostic Infectious Diseases Testing Outside Clinics: A Global Systematic Review and Meta-analysis. Open Forum Infect Dis. 2020;7(10):ofaa360.

15. Wang C, Cheng W, Li C, Tang W, Ong JJ, Smith MK, et al. Syphilis Self-testing: A Nationwide Pragmatic Study Among Men Who Have Sex With Men in China. Clin Infect Dis. 2020;70(10):2178–86.

16. World Health Organization. Recommendations and guidance on hepatitis C virus self-testing: web annex D: values and preferences on hepatitis C virus self-testing. Geneva: World Health Organization; 2021 2021.

17. Nguyen LT, Nguyen VTT, Le Ai KA, Truong MB, Tran TTM, Jamil MS, et al. Acceptability and Usability of HCV Self-Testing in High Risk Populations in Vietnam. Diagnostics (Basel). 2021;11(2).

18. Osayomi T, Adeleke R, Taiwo OJ, Gbadegesin AS, Fatayo OC, Akpoterai LE, et al. Cross-national variations in COVID-19 outbreak in West Africa: Where does Nigeria stand in the pandemic? Spatial Information Research. 2020:1–9.

19. Hamilton A, Thompson N, Choko AT, Hlongwa M, Jolly P, Korte JE, et al. HIV Self-Testing Uptake and Intervention Strategies Among Men in Sub-Saharan Africa: A Systematic Review. Front Public Health. 2021;9:594298.

20. Iliyasu Z, Kassim RB, Iliyasu BZ, Amole TG, Nass NS, Marryshow SE, et al. Acceptability and correlates of HIV self-testing among university students in northern Nigeria. Int J STD AIDS. 2020;31(9):820–31.

21. Brown B, Folayan MO, Imosili A, Durueke F, Amuamuziam A. HIV self-testing in Nigeria: public opinions and perspectives. Glob Public Health. 2015;10(3):354–65.

22. Population Council. Feasibility and acceptability of HIV self-testing among men who have sex with men in Nigeria 2018 [Available from: https://www.popcouncil.org/uploads/pdfs/2018HIV_SelfTestingMSMNigeria.pdf.

23. Jegede AS, Oshiname FO, Sanou AK, Nsungwa-Sabiiti J, Ajayi IO, Siribié M et al. Assessing Acceptability of a Diagnostic and Malaria Treatment Package Delivered by Community Health Workers in Malaria-Endemic Settings of Burkina Faso, Nigeria, and Uganda. Clin Infect Dis. 2016;63(suppl 5):S306–s11.

24. Al-Mustapha AI, Tijani AA, Oyewo M, Ibrahim A, Elelu N, Ogundijo OA, et al. Nigeria’s race to zero COVID-19 cases: True disease burden or testing failure? J Glob Health. 2021;11:03094.

25. World Health Organization. Social stigma threatens COVID-19 response but patients heal faster with everyone’s support Geneva: World Health Organization; 2020 [Available from: https://www.afro.who.int/news/social-stigma-threatens-covid-19-response-patients-heal-faster-everyones-support.

26. Shilton S, Ivanova Reipold E, Roca Álvarez A, Martínez-Pérez GZ. Assessing Values and Preferences Toward SARS-CoV-2 Self-testing Among the General Population and Their Representatives, Health Care Personnel, and Decision-Makers: Protocol for a Multicountry Mixed Methods Study. JMIR Res Protoc. 2021;10(11):e33088.

27. Ahonkhai AA, Regan S, Idigbe I, Adeniyi O, Aliyu MH, Okonkwo P, et al. The impact of user fees on uptake of HIV services and adherence to HIV treatment: Findings from a large HIV program in Nigeria. PLoS One. 2020;15(10):e0238720.

28. Ofoli JNT, Ashau-Oladipo T, Hati SS, Ati L, Ede V. Preventive healthcare uptake in private hospitals in Nigeria: a cross-sectional survey (Nisa premier hospital). BMC Health Serv Res. 2020;20(1):273.

29. Yamanis TJ, Dervisevic E, Mulawa M, Conserve DF, Barrington C, Kajula LJ, et al. Social Network Influence on HIV Testing Among Urban Men in Tanzania. AIDS Behav. 2017;21(4):1171–82.

30. Kwaghe AV, Ilesanmi OS, Amede PO, Okediran JO, Utulu R, Balogun MS. Stigmatization, psychological and emotional trauma among frontline health care workers treated for COVID-19 in Lagos State, Nigeria: a qualitative study. BMC Health Serv Res. 2021;21(1):855.

31. Olu-Owolabi FE, Amoo E, Samuel O, Oyeyemi A, Adejumo G. Female-dominated informal labour sector and family (in) stability: The interface between reproduction and production. Cogent Arts & Humanities. 2020;7(1):1788878.

32. Aduh U, Folayan MO, Afe A, Onyeaghala AA, Ajayi IO, Coker M, et al. Risk perception, public health interventions, and Covid-19 pandemic control in sub-saharan Africa. Journal of Public Health in Africa. 2020.

